# Averted mortality by COVID-19 vaccination in Belgium between 2021 and 2023

**DOI:** 10.1101/2025.02.06.25321208

**Authors:** Veerle Stouten, Izaak Van Evercooren, Catharina Vernemmen, Toon Braeye, Lucy Catteau, Mathieu Roelants, Matthieu Billuart, Thomas Lamot, Natalia Bustos Sierra, Naïma Hammami, Elias Vermeiren, Angel Rosas, Koen Blot, Anna I. Schmelz, Léonore Nasiadka, Serge Nganda, Joris A.F. van Loenhout

## Abstract

**Background:** Vaccination campaigns were rolled out primarily to limit the impact of COVID-19 on severe health outcomes, including mortality.

**Aim:** We aimed to estimate the number of averted deaths by COVID-19 vaccination in the Belgian population aged 65 years and older, between January 2021 and January 2023.

**Methods:** Nationwide data on COVID-19 infections, vaccine administrations and all-cause mortality were individually linked. We estimated Vaccine Effectiveness against COVID-19 mortality (VE) among persons having received a vaccine dose in the last 6 months, using a Cox proportional hazards model adjusted for age, sex, time since vaccination, previous infection, underlying health conditions, province and income. COVID-19 death was defined as a person with a laboratory-confirmed SARS-CoV-2 infection who died within a specified interval. Based on obtained VE estimates, vaccine coverage and national COVID-19 mortality data, we estimated the number of averted deaths.

**Results:** We estimated VE (0-59 days after vaccination) for 65-79 year and ≥80 year-olds respectively, at 81.9% (78.1%-85.1%) and 74.7% (71.2%-77.7%) during Alpha, at 90.5% (88.8%-91.9%) and 91.4% (90.4%- 92.4%) during Delta and at 84.0% (81.8%-85.9%) and 74.5% (72.4%-76.5%) during Omicron period. Among the Belgian population aged 65 years and older, we estimated 12,806 deaths averted (11,633- 13,982), representing a 54% reduction (51%-56%) in the expected deaths (without vaccination). During the Delta period COVID-19 deaths were reduced by 68%, during Omicron by 54% and during Alpha by 31%.

**Discussion:** Vaccinating against COVID-19 reduced deaths by 54% among the Belgian population aged 65 years and older, underscoring the importance of COVID-19 vaccines in reducing mortality.

## INTRODUCTION

Since the beginning of the pandemic, as of 15 September 2024, over 7 million deaths have been reported globally due to the Coronavirus disease (COVID-19) (1). Older age groups have consistently been at higher risk for severe outcomes including death (2–5). Based on a global review of publicly available data from 2020 to 2022, it was found that over 80% of all COVID-19 related deaths were among those aged ≥60 years (2). Within Belgium, according to the epidemiological COVID-19 mortality surveillance, individuals aged 65 years and above accounted for up to 92% of all COVID-19 deaths (6).

Vaccines against COVID-19 have been demonstrated to be effective in protecting against severe outcomes of the disease (7–9). Several vaccination campaigns have been rolled out in Belgium since January 2021 which were aimed at specific target groups, as well as the overall population, in successive phases. By January 1, 2023, 95.5% of the Belgian population of 65 years and older had received a primary COVID-19 vaccination scheme (10). More recent booster campaigns have focused on individuals with a higher risk of developing severe COVID-19, including the overall population of 65 years and older.

Several studies have estimated the number of COVID-19 deaths averted by vaccination in specific countries, at various stages following the introduction of COVID-19 vaccines (11–18). Of these, only few estimated the number of averted deaths also beyond 2021, during the Variant of Concern (VOC) Omicron period (17,18). One recent study demonstrated that, based on data from 2020 till 2023 provided by 54 European countries, including Belgium, over 1.4 million lives have been saved directly by COVID-19 vaccination (18). None of these studies have estimated real-life vaccine effectiveness (VE) against mortality within the same study cohort in order to calculate the number of COVID-19 averted deaths, while considering waning of protection, variation of VE by age, and hybrid immunity.

We aimed to estimate how many COVID-19 deaths have been averted in the Belgian population aged 65 years and older by COVID-19 vaccination since the beginning of the vaccination campaign (January 2021) until January 2023. First, we estimated VE against COVID-19 mortality using a population-wide cohort of ≥65 year olds, accounting for waning effectiveness of vaccination and for prior infection, and stratifying by age group and VOC-predominant period. Secondly, by combining the VE estimates with vaccine coverage and the number of reported COVID-19 deaths from the epidemiological surveillance over time, we aimed to estimate the number of COVID-19 averted deaths among Belgian inhabitants aged 65 years and older.

## METHODS

### Data sources

The LINK-VACC project was started by Sciensano, the Belgian institute of Public Health, for post-authorisation surveillance of COVID-19 vaccines (19). Datasets from national registries were linked for all individuals tested for and/or vaccinated against COVID-19: 1) the COVID-19 HealthData database, containing all performed COVID-19 laboratory tests with information on COVID-19 symptoms, 2) Vaccinnet+, the national COVID-19 vaccination registry, 3) DEMOBEL database, containing the month and year of all-cause deaths of Belgian residents, and household income, 4) Intermutualist Agency registry, containing data on reimbursed healthcare and medicines, providing information on underlying health conditions associated with a higher risk of severe COVID-19 (20), and 5) Clinical Hospital Surveillance (CHS), containing non-exhaustive data on patients hospitalized for COVID-19.

Additionally, we used data from Sciensano’s epidemiological COVID-19 mortality surveillance (21,22,6,23). This database contains COVID-19 deaths from laboratory-confirmed cases, radiologically-confirmed cases and possible cases, with demographic information and the place of death (hospitals / long-term care facilities / at home), as reported by general practitioners, since the beginning of the pandemic. As these data were collected through a separate data stream, they could not be linked to the aforementioned LINK-VACC registries.

### Study period

We used data for the period from 26 January 2021 until 31 January 2023. Periods of VOC predominant circulation were estimated using Belgium’s genomic surveillance of COVID-19 with a baseline random sampling approach (10,24). VOC predominance was defined as periods during which ≥50% of sequenced samples were identified as a certain VOC: Alpha from 26 January 2021 to 30 June 2021, Delta from 1 July 2021 to 2 January 2022 and Omicron from 3 January 2022 to 31 January 2023 (including all subvariants).

### Vaccine effectiveness against mortality

To estimate vaccine effectiveness (VE) against COVID-19 mortality among the elderly, we used a population cohort comprising all individuals of 65 years and older, based on data from LINK-VACC. Since we lacked specific information on COVID-19 as a cause of death within LINK-VACC, we defined a proxy to identify COVID-19 deaths among all-cause deaths, for the analyses to estimate VE. A COVID-19 death was defined as a person with a laboratory-confirmed SARS-CoV-2 infection who died within a specified interval thereafter. Based on previous research, we determined that the majority of COVID-19 deaths occurred within 28-33 days after a positive test (25–27). Given that only the month and year of death were available, we defined the proxy as follows: persons who tested positive in the first week of the month and died within the same month, or who tested positive in the subsequent weeks of the month and died within the same or the following month, were considered as COVID-19 deaths in our VE analyses.

We compared the number of COVID-19 deaths identified using the proxy to the number of COVID-19 deaths reported through the epidemiological surveillance system, stratifying by VOC period and by demographical characteristics. To study the impact of the potential overestimation of COVID-19 deaths by the proxy on VE analyses, we performed a first sensitivity analysis by modifying the proxy. Herein, we identified a COVID-19 death as a person with a laboratory-confirmed SARS-CoV-2 infection and who had COVID-19 symptoms or was hospitalized for COVID-19 (as registered in the CHS), and who died within the interval specified above.

To account for the time needed to develop immunity, individuals who were in the 14-day period after vaccine administration were not included in the VE analyses. Since VE against severe outcomes declines significantly over 6 months after vaccination, as shown by data from six European countries (7), we compared two groups: the ‘recently vaccinated’ comprising individuals who had received a COVID-19 vaccine dose within the past six months (<180 days) and the ‘not recently vaccinated’, comprising individuals vaccinated ≥ 180 days ago or not vaccinated at all. The ‘recently vaccinated’ category was further subdivided into 3 groups based on the time intervals since achieving the vaccination status (0-59, 60-119 and 120-179 days).

In a second sensitivity analysis, we excluded individuals who received only one dose of a primary scheme (‘partially vaccinated’) from the ‘recently vaccinated’ category and classified them as a separate vaccination status. In a third sensitivity analysis, we categorized vaccination status by also considering whether individuals had a previously registered SARS-CoV-2 infection at any time before the start of each VOC-period (‘recently vaccinated’ and ‘previous infection’ / ‘recently vaccinated’ with ‘no previous infection’ / ‘not recently vaccinated’). We did not categorize vaccination status by vaccine type or brand (10).

We estimated VE against COVID-19 mortality stratified by VOC period and by age group (65-79 and ≥80 year olds) using Cox proportional hazard models to estimate hazard ratios (HR) and 95% confidence intervals (CI). Within each VOC period and age group, the time to COVID-19 death was assessed with censoring at the earliest date of either a change in vaccination status, death from another cause, or the end of the observation period. Models incorporated the vaccination status, and as covariates age (per five years), previous laboratory-confirmed SARS-CoV-2 infection, underlying health conditions (being immunocompromised / having ≥1 other underlying health condition associated with a higher risk of severe COVID-19 / having none), province of residence and household income (deciles). VE was calculated as (1-HR) × 100%.

### Estimation of averted deaths

To estimate the number of COVID-19 deaths averted by COVID-19 vaccination in Belgium, we used three parameters: ‘reported deaths’, ‘vaccine coverage’, and ‘VE against COVID-19 mortality’, based on adapted methods developed by Machado *et al*.(28). 1) Weekly ‘reported deaths’ were derived from the national epidemiological surveillance of COVID-19 mortality, which we consider to provide the most reliable overview of COVID-19 deaths in Belgium. To adjust for potential delays in the reporting of COVID-19 deaths, we calculated the moving average of three consecutive weeks (the week of relevance, the previous week, and the following week). 2) Vaccine coverage was calculated for the Belgian population per age group, per day and aggregated per week. 3) VE against COVID-19 mortality was estimated as described above, stratified by age group and VOC period, and per 60-day interval since protection achieved by vaccination.

The weekly and cumulative number of averted COVID-19 deaths and the expected COVID-19 deaths in the absence of vaccination (the sum of reported deaths and averted deaths) were calculated by age group and by VOC period.

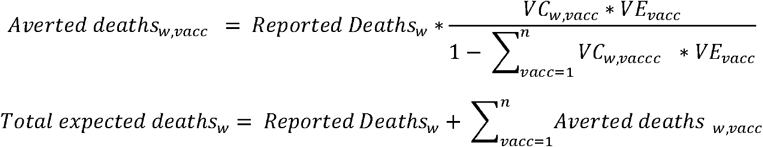

Where, *Repurted Deaths*_*w*_ = the number of recorded COVID-19 deaths in a specific week, based on date of death. *w*= the week from which the data is taken. *vacc* = vaccination statuses based on time since most recent vaccination.*VC*,_w*vacc*_ = Vaccine Coverage per week and vaccination status. *VE*_*vacc*_ = vaccine effectiveness per vaccination status.

For the reported and expected COVID-19 deaths we calculated the COVID-19 mortality rate per 100,000 individuals by week, using the weekly population size within our dataset as denominator. These weekly rates were then averaged over each VOC-period and age group. We additionally present the percentage change in expected COVID-19 mortality rate compared to the reported COVID-19 mortality rate. All analyses were performed in R version 4.2.3.

## RESULTS

### Reported deaths and vaccine coverage

Between 26 January 2021 and 31 January 2023 a total of 11,033 COVID-19 deaths have been reported through the national epidemiological surveillance among the population aged 65 years and older in Belgium (approximately 2.3 million people (29)), resulting in a weekly COVID-19 mortality rate of 4.6 per 100,000 individuals (**Table 1 and 2**). Over time, this weekly rate was higher among individuals aged ≥80 years compared to those aged 65-79 years (**Figure 1A**). The weekly COVID-19 mortality rate per 100,000 individuals, averaged by VOC-predominant period, was higher during the Alpha period (7.3) followed by the Delta (4.4) and Omicron (3.5) periods (**Table 2**).

**Table 1.** Number of reported, expected and averted COVID-19 deaths, calculated cumulatively and per month within each variant of concern (VOC) period, by age group, between 26 January 2021 and 31 January 2023, in Belgium.

| Age groups<br>(years) |  | COVID-19<br>deaths | Variant Of Concern period |  |  |  |
| --- | --- | --- | --- | --- | --- | --- |
|  |  |  | Alpha | Delta | Omicron | Total |
| 65-79 | Cumulatively | Reported | 1,442 | 1,130 | 1,484 | 4,056 |
|  |  | Expected | 2,046 (1,986 – 2,099) | 3,431 (3,222 – 3,639) | 3,531 (3,296 – 3,775) | 9,008 (8,504 – 9,513) |
|  |  | Averted | 604 (544 – 657) | 2,301 (2,092 – 2,509) | 2,047 (1,812 – 2,291) | 4,952 (4,448 – 5,457) |
|  | Per variant<br>Month | Reported | 288 | 188 | 114 | 169 |
|  |  | Expected | 409 (397 – 420) | 572 (537 – 606) | 272 (254 – 290) | 375 (354 – 396) |
|  |  | Averted | 121 (109 – 131) | 384 (349 – 418) | 157 (139 – 176) | 206 (185 – 227) |
| 80+ | Cumulatively | Reported | 2,251 | 1,533 | 3,193 | 6,977 |
|  |  | Expected | 3,309 (3,198 – 3,416) | 4,957 (4,711 – 5,199) | 6,565 (6,254 – 6,887) | 14,830 (14,162 – 15,502) |
|  |  | Averted | 1,058 (947 – 1,165) | 3,424 (3,178 – 3,666) | 3,372 (3,061 – 3,694) | 7,853 (7,185 – 8,525) |
|  | Per variant<br>Month | Reported | 450 | 256 | 246 | 291 |
|  |  | Expected | 662 (640 – 683) | 826 (785 – 867) | 505 (481 – 530) | 618 (590 – 646) |
|  |  | Averted | 212 (189 – 233) | 571 (530 – 611) | 259 (235 – 284) | 327 (299 – 355) |
| Total | Cumulatively | Reported | 3,693 | 2,663 | 4,677 | 11,033 |
|  |  | Expected | 5,355 (5,184 – 5,515) | 8,388 (7,933 – 8,838) | 10,095 (9,550 – 10,662) | 23,839 (22,666 – 25,015) |
|  |  | Averted | 1,662 (1,491 – 1,822) | 5,725 (5,270 – 6,175) | 5,418 (4,873 – 5,985) | 12,806 (11,633 – 13,982) |
|  | Per variant<br>Month | Reported | 739 | 444 | 360 | 460 |
|  |  | Expected | 1,071 (1,037 – 1,103) | 1,398 (1,322 – 1,473) | 777 (735 – 820) | 993 (944 – 1042) |
|  |  | Averted | 332 (298 – 364) | 954 (878 – 1029) | 417 (375 – 460) | 534 (485 – 583) |

Reported: COVID-19 deaths recorded in the epidemiological COVID-19 mortality surveillance; Averted: COVID-19 deaths averted by vaccination, estimated based on adapted methods developed by Machado et al, by using the reported deaths, vaccine coverage and VE against COVID-19 mortality; Expected: COVID-19 deaths expected without vaccination which is the sum of the reported and averted deaths. Numbers between brackets present the 95% CI of expected and averted deaths resulting from the 95% CI of the VE estimates. Data are by age group and by Variant Of Concern (VOC) periods for Belgium: the Alpha period from 26 January 2021 to 30 June 2021, the Delta period from 1 July 2021 to 2 January 2022 and the Omicron period from 3 January 2022 to 31 January 2023, and for the entire study period. Per variant month: The cumulative number of deaths for each VOC period was divided by the number of months within each VOC period.

**Table 2.** Weekly COVID-19 mortality rates per 100,000 individuals (and the 95% confidence interval) of reported and expected COVID-19 deaths, and the percentage reduction between these, by age group and by VOC-period, between 26 January 2021 and 31 January 2023 in Belgium.

| Age groups<br>(years) | COVID-19<br>mortality rates<br>per 100,000<br>individuals | Variant Of Concern period |  |  |  |
| --- | --- | --- | --- | --- | --- |
|  |  | Alpha | Delta | Omicron | Total |
| 65-79 | Reported | 4.1 | 2.6 | 1.6 | 2.4 |
|  | Expected | 5.8 (5.6 – 5.9) | 8.0 (7.5 – 8.5) | 3.8 (3.5 – 4.0) | 5.2 (5.0 – 5.5) |
|  | % Reduction | 29.6 (27.4 – 31.4) | 67.1 (64.9 – 68.9) | 57.9 (54.9 – 60.6) | 55.0 (52.3 – 57.4) |
| 80+ | Reported | 15.2 | 8.9 | 8.4 | 9.9 |
|  | Expected | 22.3 (21.6 – 23.1) | 28.8 (27.4 – 30.2) | 17.2 (16.4 – 18.1) | 21.3 (20.3 – 22.2) |
|  | % Reduction | 32.2 (29.8 – 34.3) | 69.1 (67.5 – 70.6) | 51.2 (48.8 – 53.4) | 53.2 (51.0 – 55.3) |
| Total | Reported | 7.3 | 4.4 | 3.5 | 4.6 |
|  | Expected | 10.7 (10.3 – 11.0) | 13.9 (13.2 – 14.7) | 7.6 (7.2 – 8.1) | 9.9 (9.4 – 10.3) |
|  | % Reduction | 31.1 (28.8 – 33.1) | 68.3 (66.4 – 69.9) | 53.6 (50.9 – 56.0) | 53.8 (51.4 – 56.0) |

**Figure 1.**
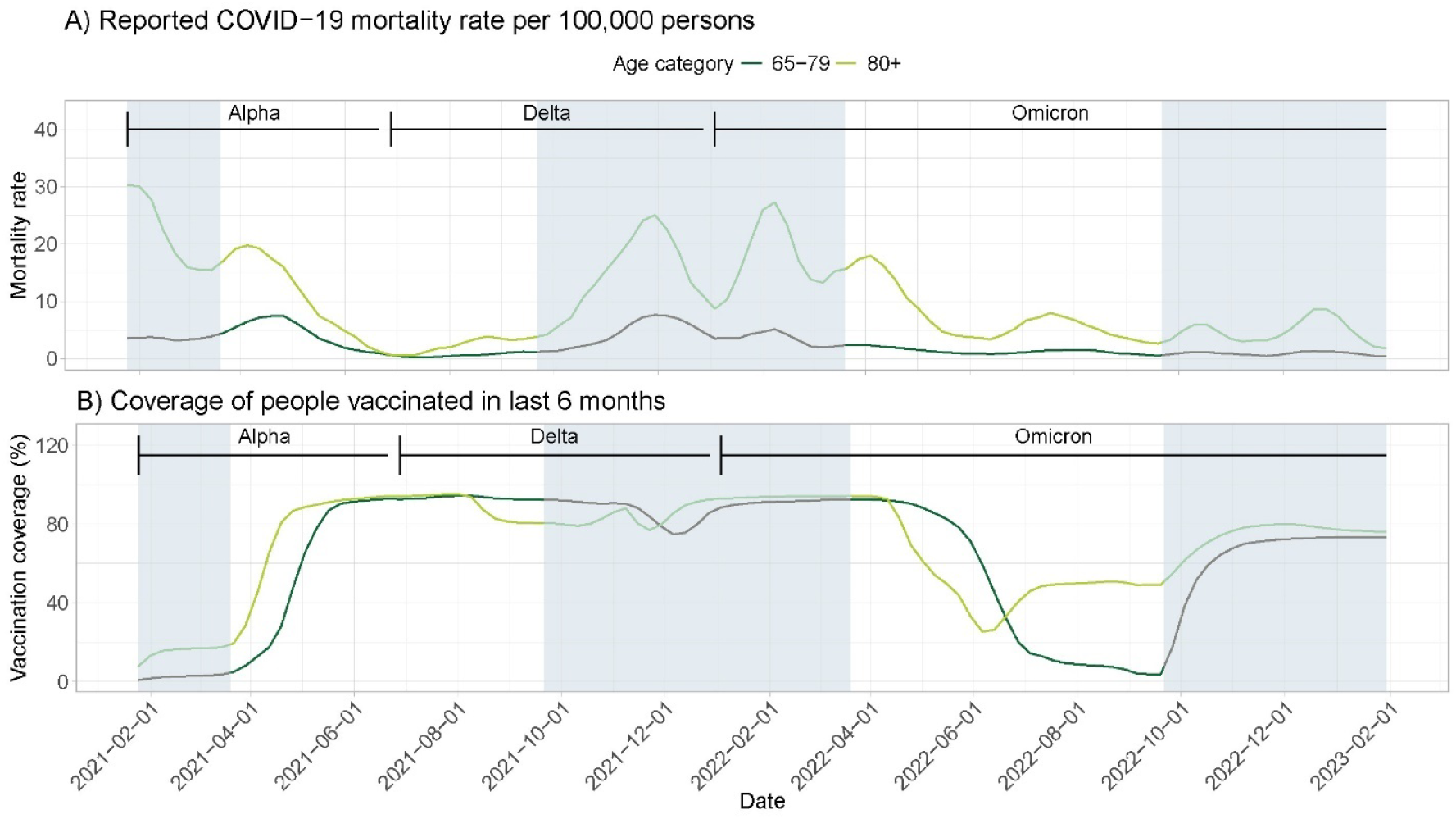
**A**. Reported weekly COVID-19 mortality rates per 100,000 individuals over time, by VOC predominant period and age groups (65-79 years and ≥80 years) in Belgium (26 January 2021 – 31 January 2023), based on the national epidemiological COVID-19 mortality surveillance. **B**. Vaccine coverage (percentage of individuals vaccinated with a dose of a COVID-19 vaccine in the last 6 months) over time, by VOC predominant period and age groups (65-79 years and ≥80 years) in Belgium (26 January 2021 – 31 January 2023). Grey areas indicate autumn and winter periods (from 21st of September to 21st of March).

Since January 2021, the population aged 65 years and older residing in Belgium has been regularly invited to receive a COVID-19 vaccine dose. Following each vaccination campaign, the vaccine coverage - the percentage of people who received a COVID-19 vaccine in the last six months - rose to at least 70%, for both age groups, during autumn and winter periods, when higher transmission of SARS-CoV-2 was expected (**Figure 1B**) (10,30).

### Vaccine effectiveness against COVID-19 mortality

The number of COVID-19 deaths identified using our proxy was, compared to the number reported through the epidemiological surveillance, similar in the alpha period, slightly higher in the Delta period (1.5 times higher), and significantly higher during the Omicron period (2.8 times higher) (**Supplement 1**). However, COVID-19 deaths identified by either method were comparable in terms of demographics (age, gender, geographical region).

In general, VE among the elderly (≥80 years) was lower compared to those aged 65-79 years. During the Alpha period, VE at the initial time interval of 0-59 days after vaccination was estimated at 81.9% (CI 78.1%-85.1%) for those aged 65-79 years and at 74.7% (CI 71.2%-77.7%) for those aged ≥80 years (**Figure 2A**). During the Delta period, the initial VE was 90.5% (CI 88.8%-91.9%) for those aged 65-79 years, and 91.4% (CI 90.4%-92.4%) for those aged ≥80 years (**Figure 2B**). When Omicron was predominant, initial VE was estimated at 84.0% (CI 81.8%-85.9%) for those aged 65-79 years, and at 74.5% (CI 72.4%-76.5%) for those aged ≥80 years (**Figure 2C**). We observed a weekly waning rate in VE estimates of on average 1.2% for the entire study period, and more specifically of -0.6%, 1.0% and 2.2% within the Alpha, Delta and Omicron periods respectively.

**Figure 2:**
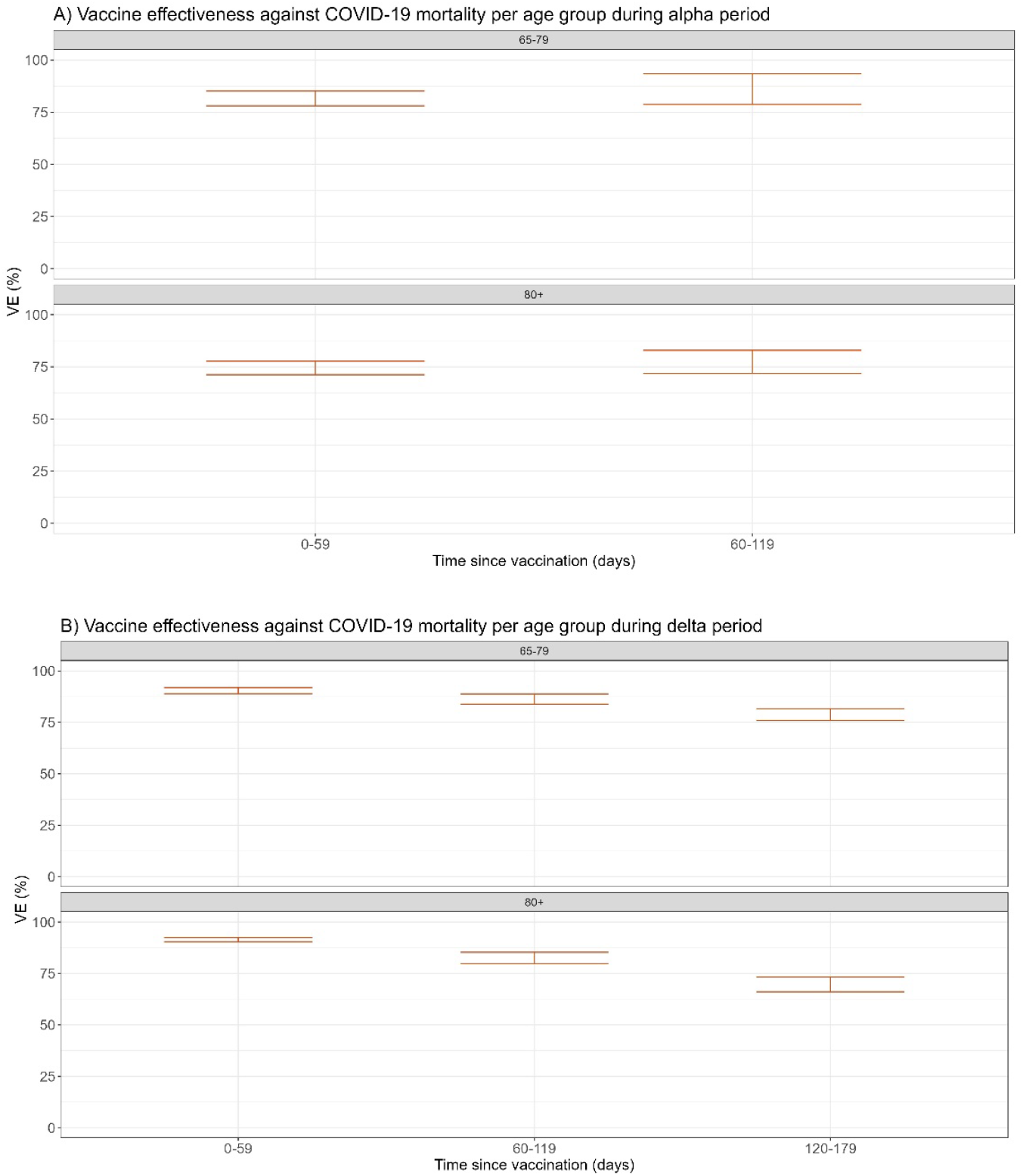
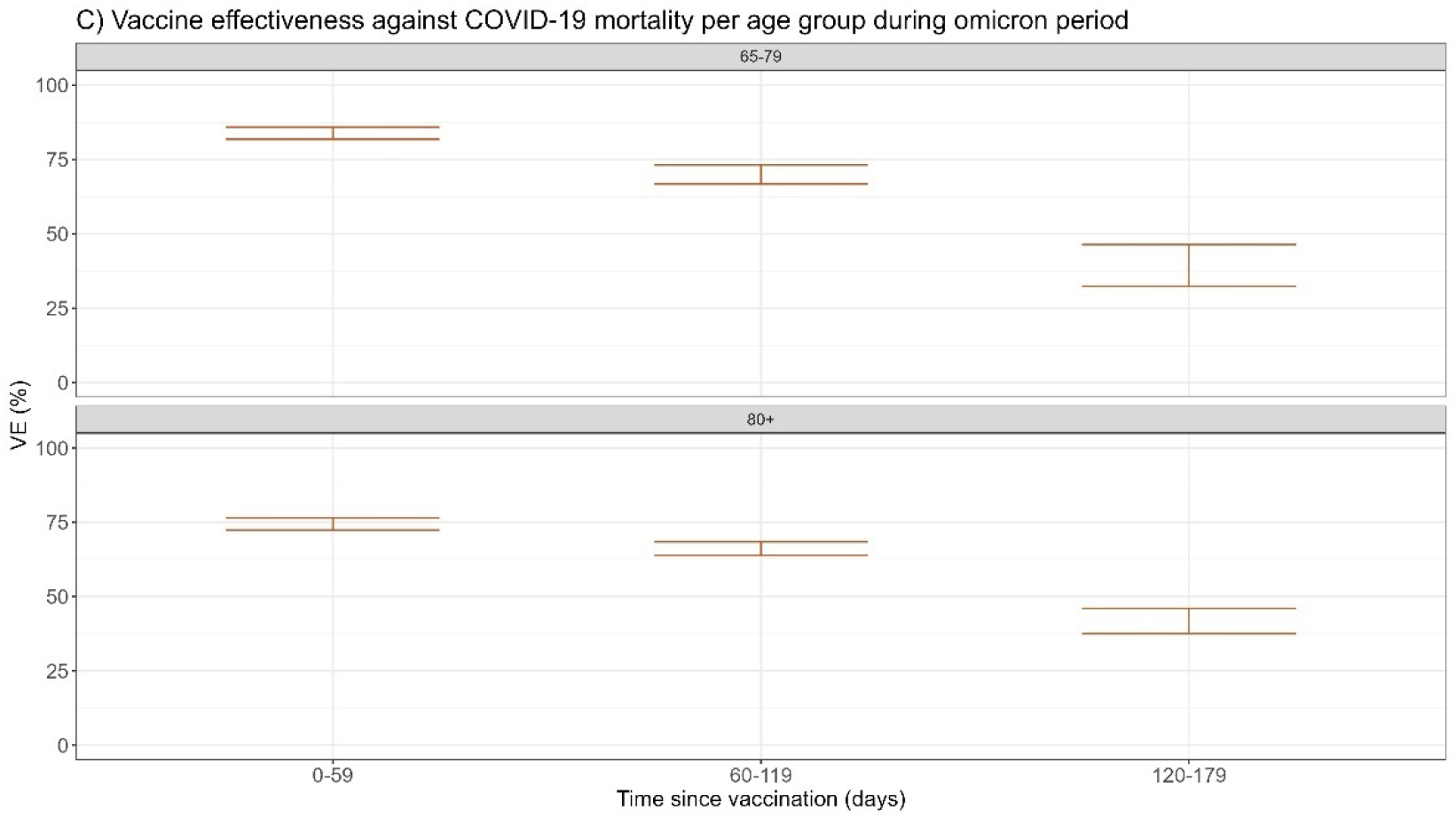
Vaccine effectiveness against COVID-19 mortality by age group and time since vaccination (60 days interval) during: **A**. the Alpha period from 26 January 2021 till 30 June 2021), **B**. the Delta period from 1 July 2021 to 2 January 2022 and **C**. the Omicron period from 3 January 2022 till 31 January 2023.

VE estimates and their 95% confidence intervals are presented for ‘recently vaccinated’ individuals (having received a COVID-19 vaccine dose within the past six months (<180 days)) with ‘non-recently vaccinated’ individuals (including unvaccinated individuals) serving as reference group.

In the first sensitivity analyses, the reported COVID-19 mortality rates based on the modified COVID-19 death proxy (deaths with reported COVID-19 symptoms and/or hospitalization) were slightly lower than those reported through the epidemiological surveillance but followed a similar trend over time (**Supplement 1**). Using the modified COVID-19 death proxy to estimate VE against mortality resulted in limited changes (an average absolute difference (AAD) of 4%) in VE estimates compared to using the initial proxy (**Supplement 2**).

In the second sensitivity analysis, excluding the ‘partially vaccinated’ from the ‘recently vaccinated’ category, the initial VE for ‘recently vaccinated’ individuals was slightly higher during the Alpha period than in the main analysis (AAD of 8%), but comparable during the Delta and Omicron period (AAD of 0.3%) (**Supplement 3**).

In the third sensitivity analysis, estimating VE against mortality separately for individuals with or without a previous registered SARS-CoV-2 infection, estimates were comparable right after vaccination (an average absolute difference of 4%). However, VE was slightly higher for those with a previous infection at the later time interval of 120-179 days after vaccination (AAD of 14%), indicating a prolonged protection by hybrid immunity **(supplement 4)**.

### Averted deaths

We estimated that COVID-19 vaccination in Belgium resulted in the averting of 12,806 COVID-19 deaths (CI 95% 11,633 to 13,982) among individuals aged 65 years and older between January 2021 and January 2023. Of these, 4,952 (39%) deaths were averted among those aged 65- 79 years, and 7,853 (61%) among those aged ≥80 years (**Table 1**). On average, the number of averted COVID-19 deaths per variant per month was higher during the Delta period compared to the Omicron and Alpha period (954 versus 417 and 332, respectively).

During the study period, the total reported number of COVID-19 deaths was 11,033. Considering the deaths averted by COVID-19 vaccination, we estimated that the expected number of COVID-19 deaths would have been more than double, namely 23,839 (22,666 – 25,015).

The overall weekly COVID-19 mortality rate per 100,000 individuals was decreased by 54% (CI 51% to 56%) over the entire study period (**Table 2**). By late 2021, COVID-19 mortality rates showed a larger difference between expected and reported deaths, reflecting the higher number of averted deaths at the end of the delta period and the beginning of the omicron period, compared to the alpha period (**Figure 3A**). Cumulative COVID-19 mortality rates increased more steeply for individuals aged ≥80 years compared to those aged 65-79 years, but the percentage reduction in expected mortality rates was comparable for those aged ≥80 years and those aged 65-79 years (53% and 51% respectively) (**Figure 3B and Table 2**).

**Figure 3:**
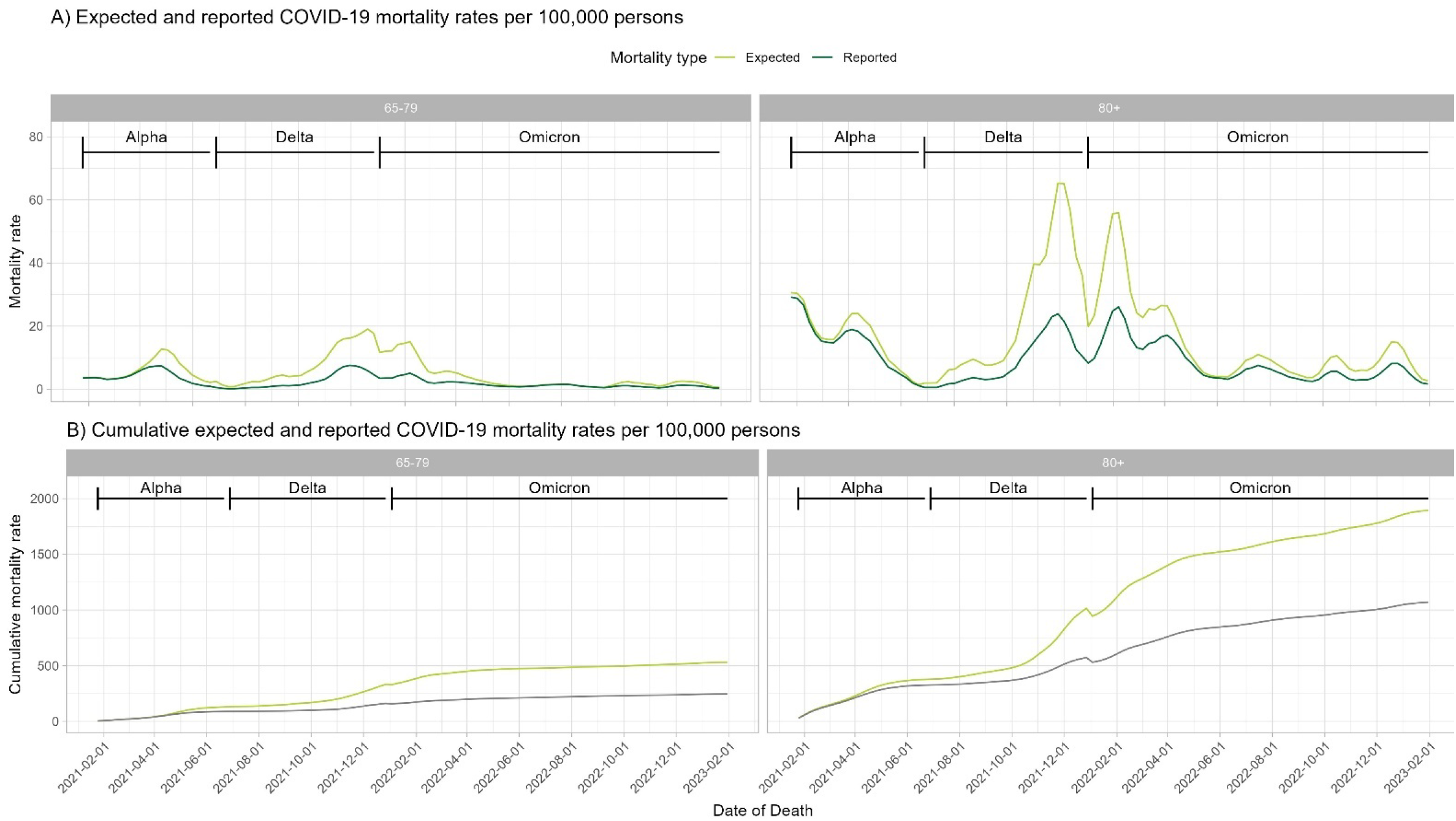
Expected and reported COVID-19 mortality rates per 100,000 individuals, by age group, and by variant of concern period between 26 January 2021 and 31 January 2023, in Belgium.

Reported: COVID-19 mortality rate based on data from the epidemiological COVID-19 mortality surveillance; Expected: COVID-19 mortality rate estimated in the absence of vaccination; % Reduction: calculated as the percentage of decrease in the expected mortality rate compared to the reported mortality rate. Numbers between brackets present the 95% CI of expected deaths resulting from the 95% CI of the VE estimates.

All calculations of expected and averted COVID-19 deaths were repeated in the three aforementioned sensitivity analyses. When using the modified proxy (only inclusion of persons with COVID-19 symptoms and/or hospital admission) to identify COVID-19 deaths, small changes in the number of deaths averted and in the percentage reduction in mortality rates were observed (0.1%-3.6%), compared to using the initial proxy (**Supplement 5 A/B**). Sensitivity analyses using VE estimates that accounted for partial vaccination or for previous infection, also resulted in small changes in the percentage reduction of mortality (0.1-1.9%) (data not shown).

## DISCUSSION

Our results demonstrate that during the first two years of the vaccination campaign, COVID-19 mortality was reduced by 54% (CI 51% to 56%), corresponding to a total of 12,806 averted deaths (CI 11,633 to 13,982) among the Belgian population aged 65 years and older. VE against COVID-19 mortality was high shortly after vaccination (75%-91%) but waned over time during the Delta and more rapidly during the Omicron period. VE estimates were generally higher among individuals aged 65-79 years compared to those aged ≥80 years. Additionally, VE was slightly higher and waned less rapidly for individuals with a previous SARS CoV-2 infection.

Our findings on VE against COVID-19 mortality align with results from other studies on VE against mortality in populations over 60 years (15,31–33) and provide additional insight to what extent VE against mortality wanes over a longer follow-up period of up to six months after protection obtained by vaccination. The results regarding averted deaths are consistent with previous studies that have examined the impact of vaccination in Europe and globally (12,16,18). Two studies on data from 54 European regions, including Belgium, demonstrated that COVID-19 vaccination programs reduced COVID-19 mortality in Europe by 51% in the first 12 months of the pandemic, and by 59% based on data covering a large period of the pandemic (12,18). However, direct comparisons with these studies are challenging due to methodological differences. These studies used different VE estimates based on meta-analyses from the literature, which could not be stratified by age-group or infection status. Furthermore, the literature-based assumed general waning rate of 0.25% per week in the latter study differed from the waning rate we observed in our study (1.2%), which also differed by VOC period: -0.6%; 1.0% and 2.2% for the Alpha, Delta and Omicron periods respectively.

We observed a relatively comparable impact among individuals aged 65-79 years and those aged ≥80 years. While individuals aged ≥80 years had higher cumulative reported and expected COVID-19 mortality rates and accounted for more than half of the total averted COVID-19 deaths in the population aged 65 years and older (61%), they also showed a slightly lower reduction in mortality rates compared to 65-79 year olds (53% versus 55%). This can be attributed to the lower VE estimates observed in individuals aged ≥80 years.

Our analyses revealed a higher impact of COVID-19 vaccination on reducing mortality during the Delta and Omicron periods compared to the Alpha period (68%, 54%, and 31% respectively). The vaccination campaign was still being rolled out during the Alpha period, resulting in a higher vaccine coverage during Delta and Omicron periods. VE against mortality waned more rapidly during the Omicron period than during Delta, consistent with other reports (7,8,34–36), contributing to a lower absolute number of averted COVID-19-deaths during Omicron than during Delta. However, during Omicron COVID-19 mortality rates were still reduced by 54%, even though infection severity had decreased during Omicron (37–40), reflecting the ongoing effectiveness of COVID-19 vaccines in protecting against severe outcomes. Our results support the recently published Strategic Advisory Group of Experts (SAGE) recommendations for COVID-19 vaccination in the context of Omicron and high population immunity (41).

Our study has several strengths. Following comparisons with all-cause mortality and death certificates, the epidemiological COVID-19 mortality surveillance was considered sensitive in capturing a large fragment of COVID-19 deaths in Belgium, due to which we have a good estimate of the real number of COVID-19 deaths over the study period (42,43). Through the LINK-VACC project, we were able to estimate VE against COVID-19 mortality in a population-wide cohort. This enabled us to study potential variations in VE by VOC-predominant period and by age group, the waning of protection and the impact of hybrid immunity in a real-life setting with country-specific estimates, while correcting for various relevant confounders. Sensitivity analyses accounting for partial vaccination or previous infection, resulted in only small changes in VE and in the percentage reduction in mortality.

Our study also has several limitations. We could not adjust for potential confounders such as antiviral or other therapies that might have an impact on COVID-19 mortality. We used a proxy based on laboratory- confirmed infections and all-cause mortality to identify COVID-19 deaths for our VE analyses, potentially overestimating true COVID-19 deaths. However, sensitivity analyses using a more specific proxy including also information on COVID-19 related symptoms and hospitalizations due to COVID-19 led to similar results in terms of VE and the number of averted deaths. Underestimation of COVID-19 deaths by the proxy was likely limited as the majority of individuals were tested before dying from COVID-19, as known based on data from the epidemiological surveillance (96% of reported COVID-19 deaths were lab-confirmed cases). Additionally, there could have been a reporting bias in the number of reported COVID- 19 deaths through the epidemiological COVID-19 mortality surveillance. This surveillance was of high quality, as examined by Sciensano by comparing COVID-19 deaths from the epidemiological surveillance with COVID-19-associated deaths from the causes of death database using death certificates: the surveillance identified 90% of COVID-19-associated deaths from death certificates in 2020 (n=19,801) and 85% in 2021 (n=10,052). Underreporting by general practitioners of individuals who died at home was likely, although most deaths occurred in hospitals or long-term care facilities where reporting was well covered. Overreporting of COVID-19 deaths through potential misclassification of the cause of death appeared to be limited, as 84% of deaths in the epidemiological surveillance also had COVID-19 indicated as the underlying cause on the death certificates (43).

In conclusion, our study estimated COVID-19 mortality to be reduced by 54% by COVID-19 vaccination among the Belgian population aged 65 years and older during the first two years of the vaccination campaign, underscoring the significant impact of COVID-19 vaccines. The findings highlight the importance of COVID-19 vaccination in reducing severe outcomes, and support ongoing vaccination efforts, including booster doses, to maintain high levels of protection against COVID-19 mortality.

## STATEMENTS

### Ethical statement

Data linkage and collection within the data-warehouse has been approved by the information security committee. The study was conducted in accordance with the Declaration of Helsinki. Ethical approval was granted for the gathering of data from hospitalised patients by the Committee for Medical Ethics from the Ghent University Hospital (reference number BC-07507) and authorisation for possible individual data linkage using the national register number from the Information Security Committee (ISC) Social Security and Health (reference number IVC/KSZG/20/384). Linkage of hospitalised patient data to vaccination, and testing within the LINK-VACC project was approved by the Medical Ethics Committee UZ Brussels– VUB on 3 February 2021 (reference number 2020/523), and authorisation from the ISC Social Security and Health (reference number IVC/KSZG/21/034).

Informed consent was waived based on art. 6 and 9 of the GDPR. The collection is allowed based on general interest (art. 6 GDPR) and regarding article 9 § 2 of the GDPR: processing is necessary for reasons of public interest in the area of public health, such as protecting against serious cross-border threats to health, or ensuring high standards of quality and safety of health care and of medicinal products or medical devices, on the basis of Union or Member State law which provides for suitable and specific measures to safeguard the rights and freedoms of the data subject, in particular, professional secrecy.

### Use of artificial intelligence tools

None declared.

### Data availability

As the LINK-VACC project is not an open-access platform, the individual level data are only available to researchers working on the project. However, general descriptive statistics from the registries used in LINK-VACC are available from https://epistat.sciensano.be/covid/. This includes, among others, the number of deaths by date, age, sex and province or the number of administered vaccines by date, region, age, sex, brand and dose.”

## Acknowledgements

We would like to thank everyone who helped in providing and processing the data to enable surveillance of COVID-19, especially those surveillances that are on a voluntary basis. We greatly appreciate your time and effort.

## Authors’ contributions

Management of the LINKVACC environment: VS, JVL, LC. (Data) management testing and vaccination registry: IVE, MB, EV. (Data) management of the epidemiological COVID-19 mortality surveillance: CV, NBS, SN. Study set-up and data analysis: VS, IVE, JVL, TB, CV, LC. Writing and reviewing the article: VS, JvL, IVC, CV, TB, LC, MR, MB, TL, NBS, NH, EV, AR, KB, AIS, LN, SN.

