## Supplementary files for "Averted mortality by COVID-19 vaccination in Belgium between 2021 and 2023"

### Supplement 1

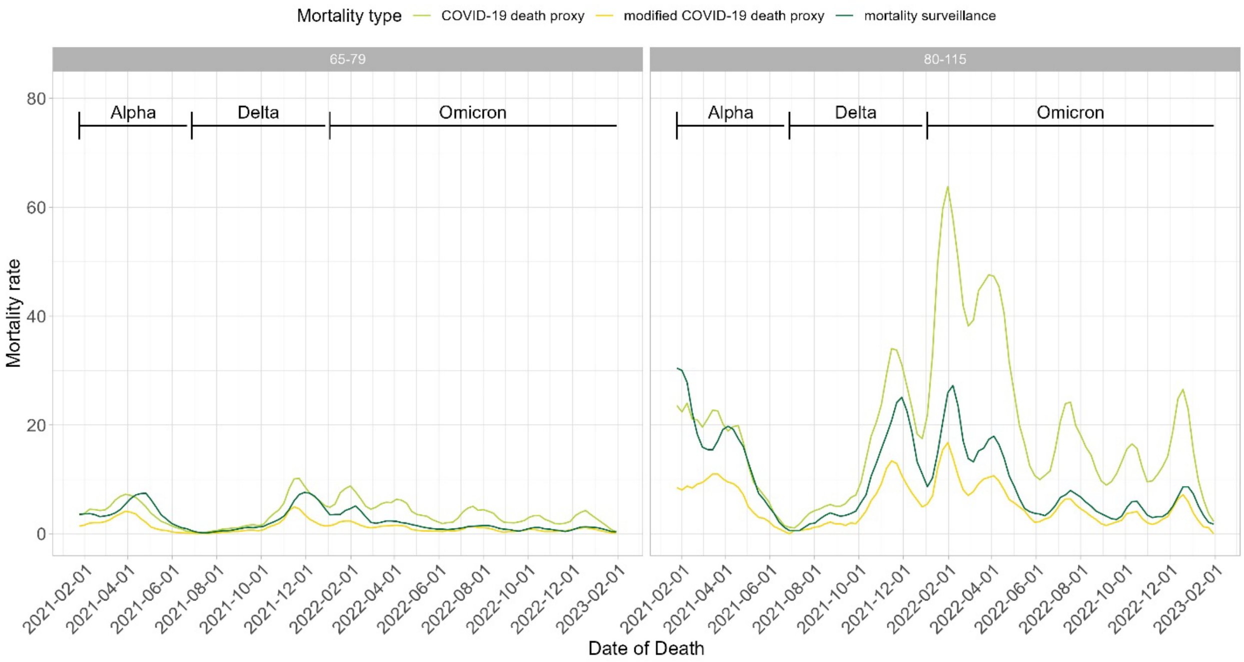


**Supplement 1.** Reported mortality rates per 100,000 persons, per identification method of COVID-19 deaths, per age group, between 2021 and 2023, in Belgium.

**COVID-19 death proxy:** A COVID-19 death was defined as a person with any laboratory-confirmed SARS-CoV-2 infection and died within a certain interval thereafter. More specifically, a person who tested positive in the first week of the month and died within the same month, or who tested positive in the subsequent weeks of the month and died within the same or the following month, were considered as COVID-19 deaths.

**Modified COVID-19 death proxy**: a COVID-19 death was identified as a person with a laboratory-confirmed SARS-CoV-2 infection, with registered COVID-19 symptoms or was hospitalized for COVID-19, who died within the same interval thereafter, as indicated above.

**Mortality surveillance:** National epidemiological surveillance of COVID-19 mortality. This database contains data on laboratory-confirmed COVID-19 deaths from three types of sources: 1) hospitals, 2) long-term care facilities, and 3) general practitioners, since the beginning of the pandemic.

#### Supplement 2

A) Vaccine effectiveness against COVID-19 mortality per age group during alpha period

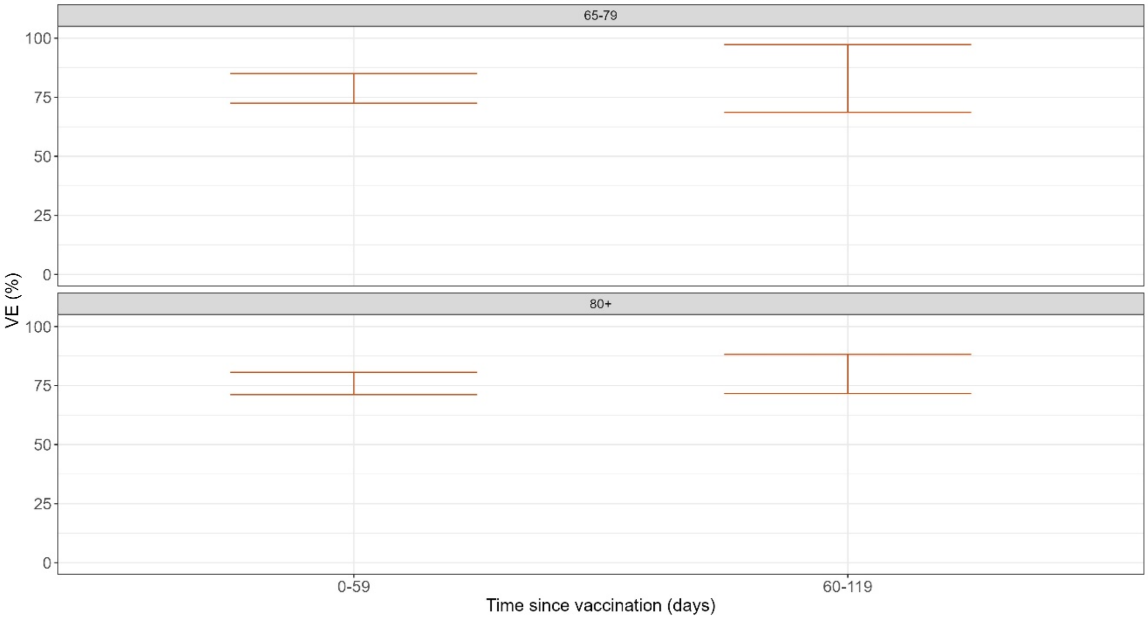


B) Vaccine effectiveness against COVID-19 mortality per age group during delta period

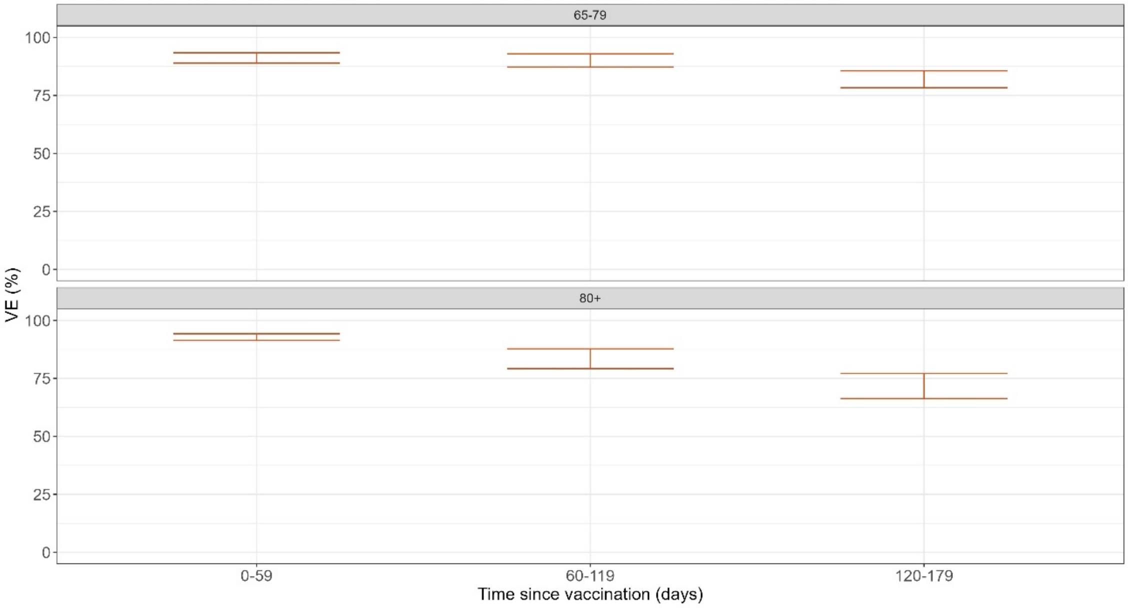

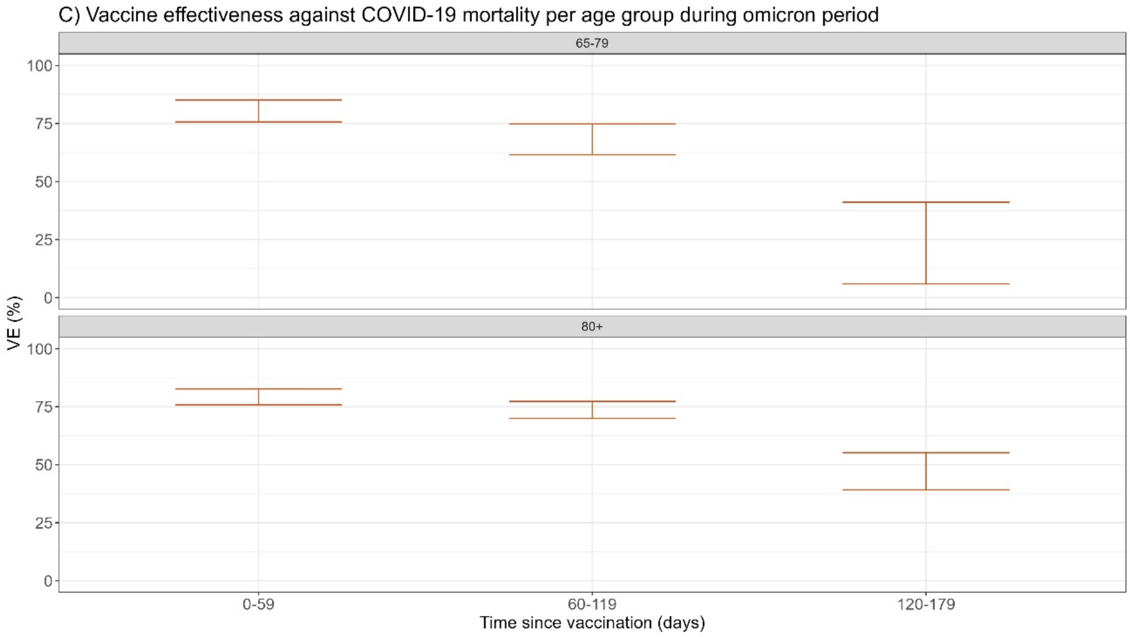


**Supplement 2:** Vaccine effectiveness against COVID-19 mortality, based on a modified proxy for COVID-19 death including information on symptoms or hospitalizations, per age group during **A.** Alpha period **B.** Delta period and **C.** Omicron period.

For these VE analyses, a COVID-19 death was identified as a person with a laboratory-confirmed SARS-CoV-2 infection, with registered COVID-19 symptoms or was hospitalized for COVID-19, who died within an interval thereafter. More specifically, a person who tested positive in the first week of the month and died within the same month, or who tested positive in the subsequent weeks of the month and died within the same or the following month, were considered as COVID-19 deaths.

VE estimates are shown for persons recently vaccinated the last six months with as reference persons not recently vaccinated (including unvaccinated persons). VE estimates with a CI >50% were not taken into account in the analyses and are not displayed.

#### Supplement 3

A) Vaccine effectiveness against COVID-19 mortality per age group during alpha period

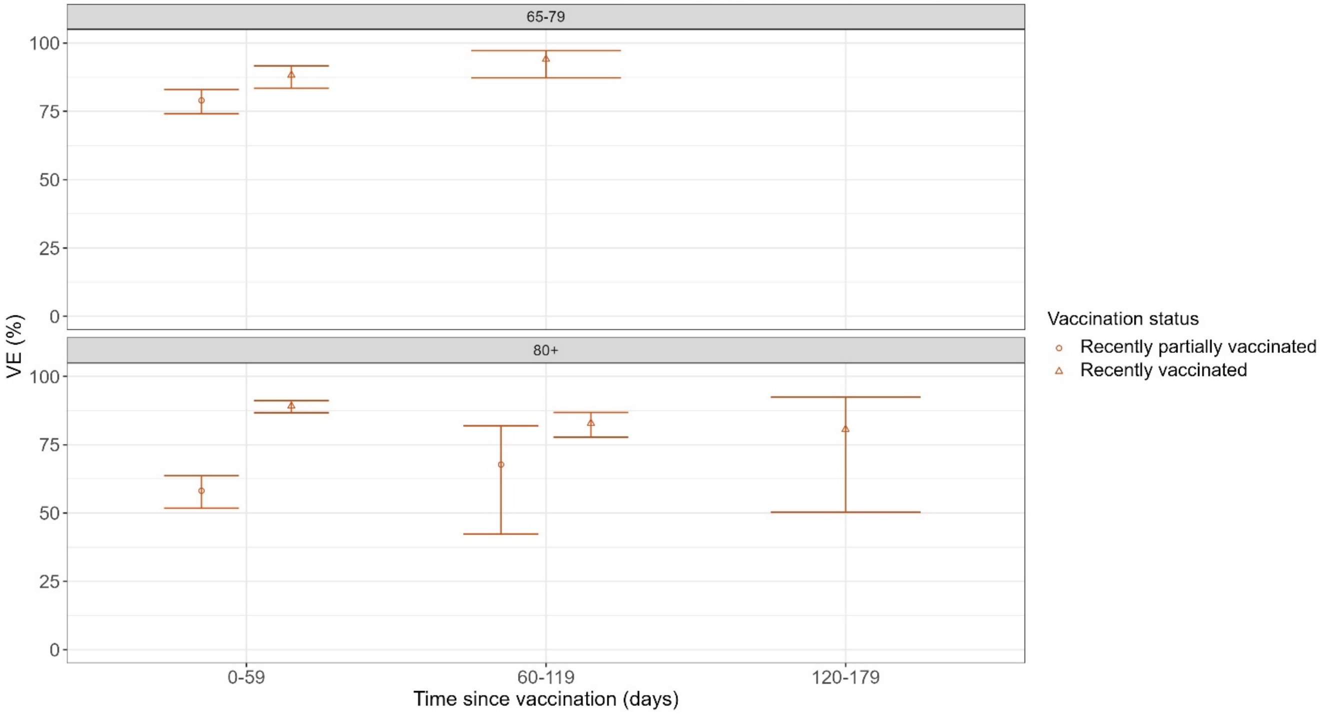


B) Vaccine effectiveness against COVID-19 mortality per age group during delta period

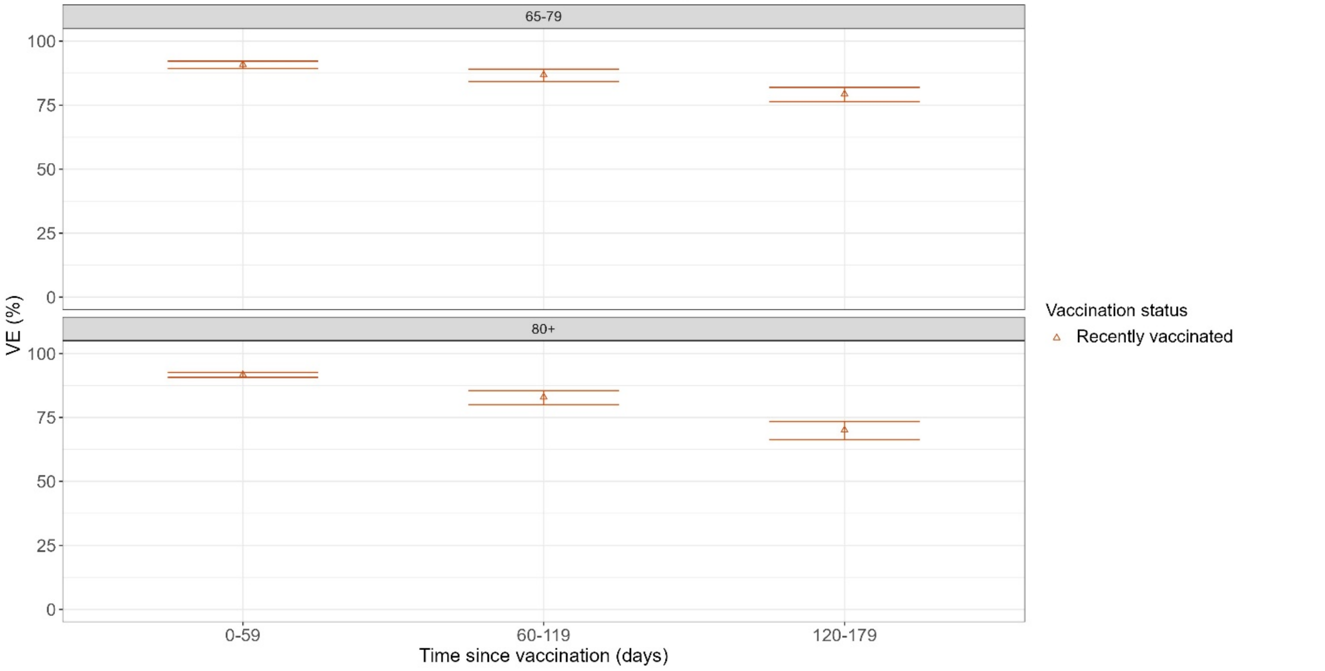

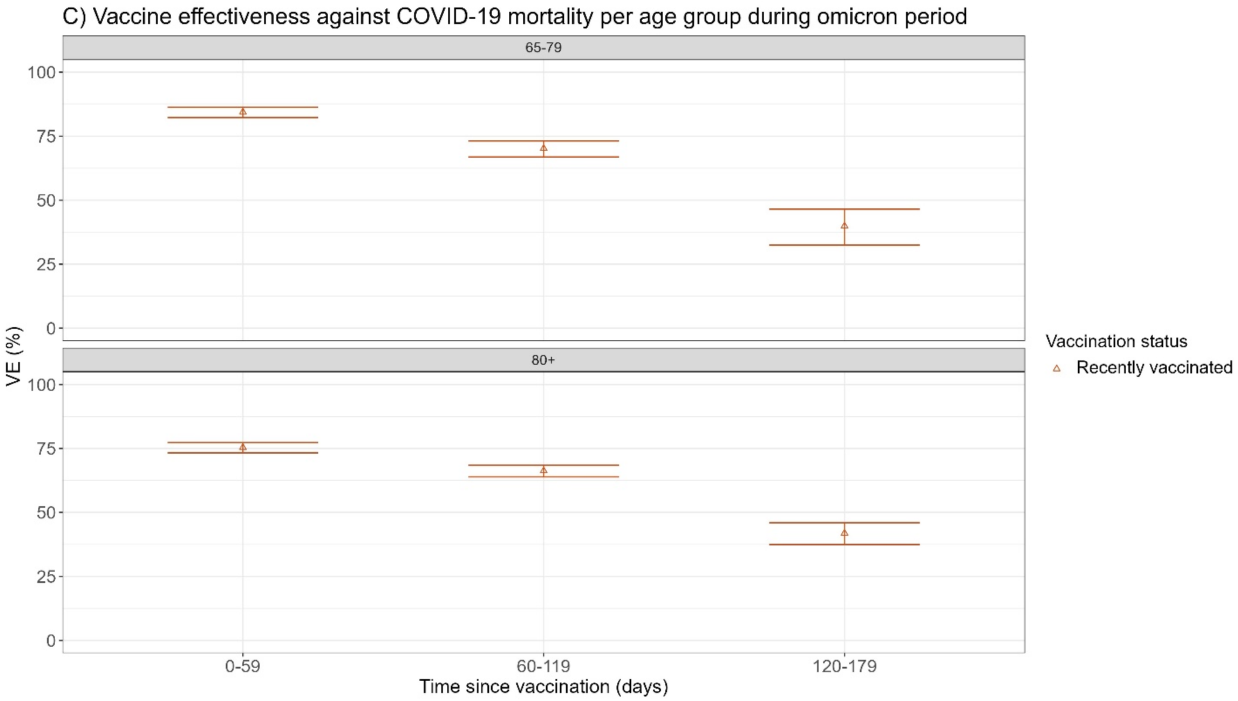


**Supplement 3:** Vaccine effectiveness against COVID-19 mortality, excluding persons who received only one dose of a primary scheme (partially vaccinated) out of the recently vaccinated, and classifying these as a separate vaccination status, per age group during **A.** Alpha period **B.** Delta period and **C.** Omicron period.

Reference group consisted of persons not recently vaccinated (including unvaccinated persons).VE estimates with a CI >50% were not taken into account in the analyses and are not displayed.

#### Supplement 4

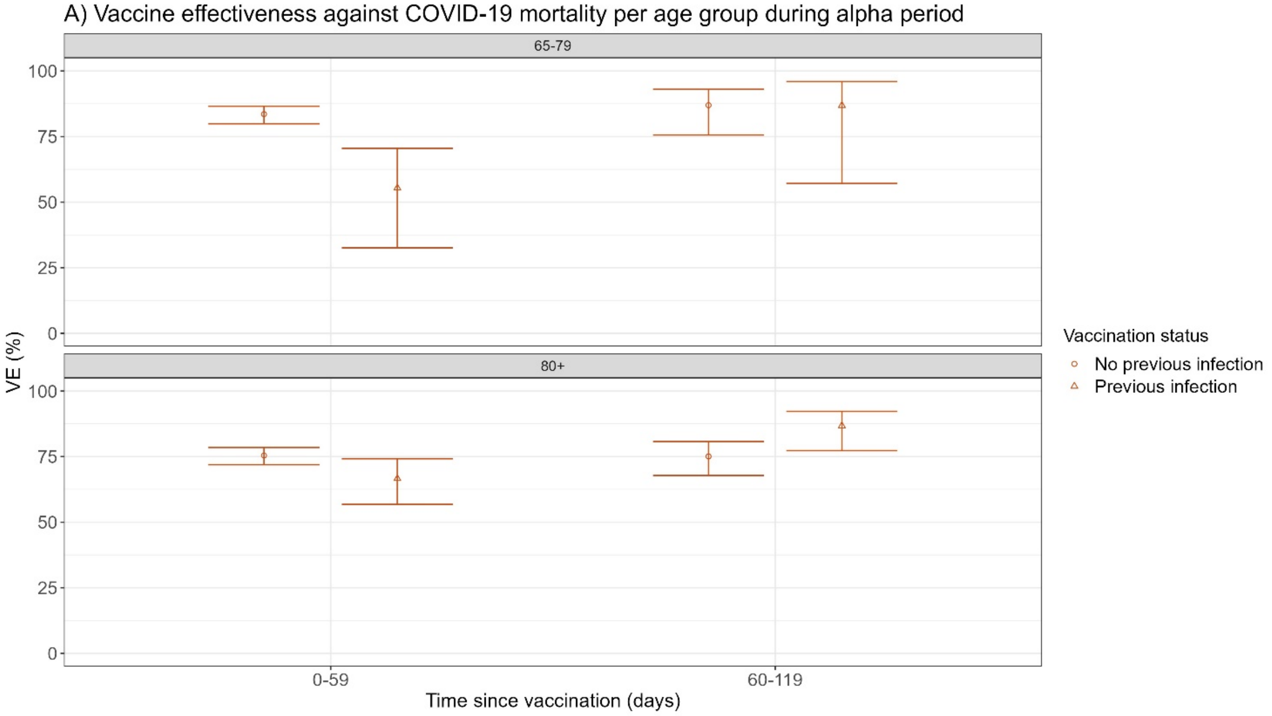


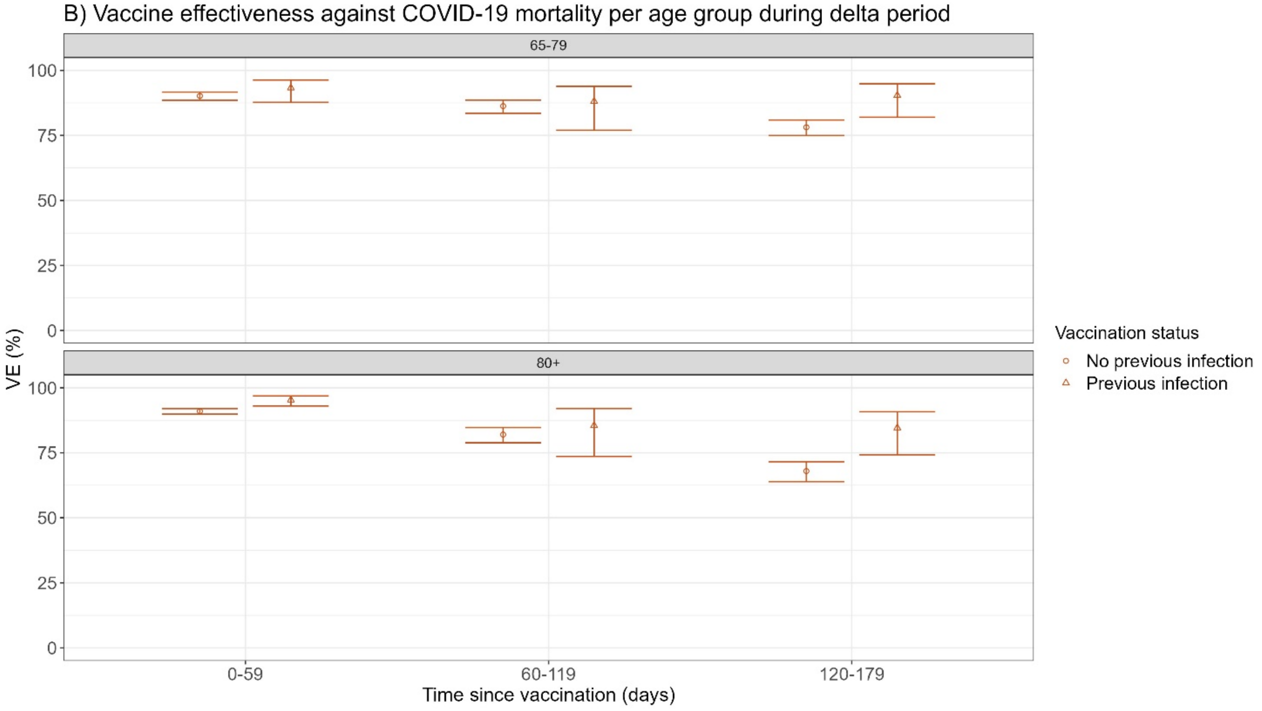

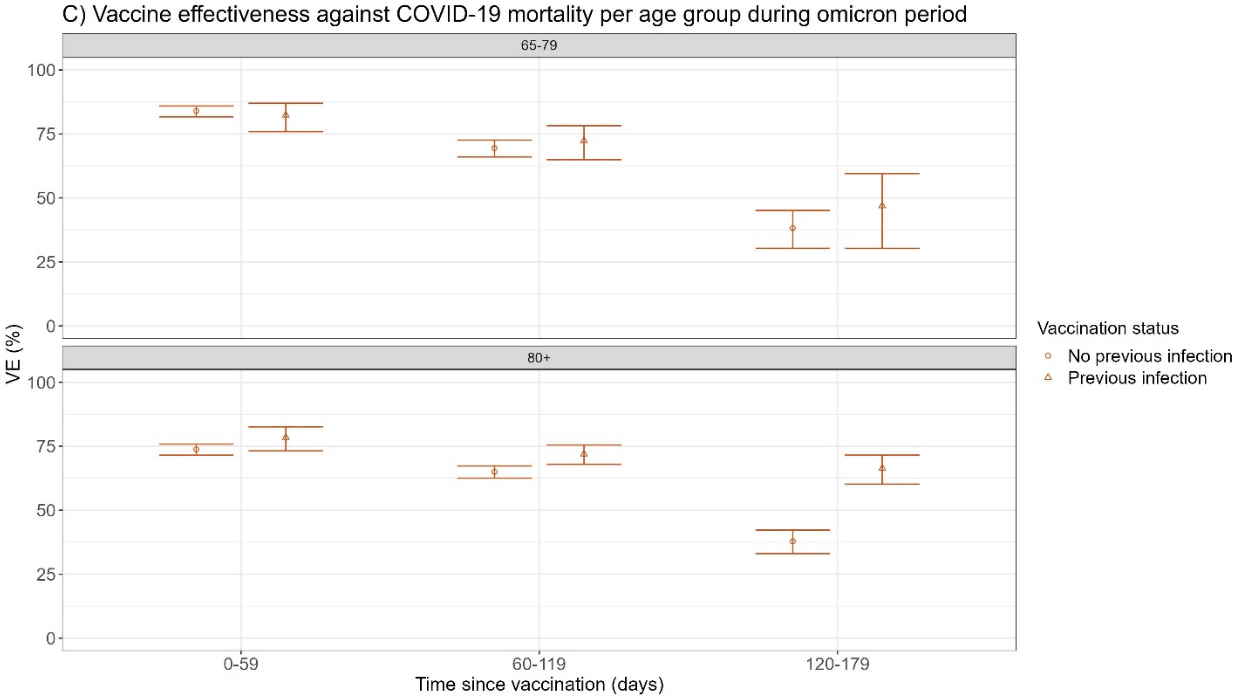


**Supplement 4:** Vaccine effectiveness against COVID-19 mortality, estimated separately for Recently Vaccinated persons who had a previous registered SARS-CoV-2 infections or not, per age group during **A.** Alpha period **B.** Delta period and **C.** Omicron period.

VE estimates are shown for persons recently vaccinated the last six months with as reference persons not recently vaccinated (including unvaccinated persons). VE estimates with a CI >50% were not taken into account in the analyses and are not displayed.

#### Supplement 5

**Supplement 5A:** Number of reported, expected and averted deaths*, calculated cumulatively and per month within a VOC-period, by age group and by VOC-period, between 26 January 2021 and 31 January 2023 in Belgium.

* using VE against mortality based on a <u>modified proxy for COVID-19 death</u> including reported symptoms or hospitalizations related to COVID-19

| Age groups in years | | Mortality | Alpha | Delta | Omicron | Total |
|---|---|---|---|---|---|---|
| 65-79 | Cumulatively | Reported | 1,442 | 1,130 | 1,484 | 4,056 |
| | | Expected | 2,017 (1,912 – 2,102) | 3,716 (3,385 – 4,035) | 3,285 (2,848 – 3,761) | 9,018 (8,145 – 9,897) |
| | | Averted | 575 (470 - 660) | 2,586 (2,255 – 2,905) | 1,801 (1,364 – 2,277) | 4,962 (4,089 – 5,841) |
| | Per variant Month | Reported | 288 | 188 | 114 | 169 |
| | | Expected | 403 (382 - 420) | 619 (564 - 672) | 253 (219 - 289) | 376 (339 - 412) |
| | | Averted | 115 (94 - 132) | 431 (376 - 484) | 139 (105 - 175) | 207 (170 - 243) |
| 80+ | Cumulatively | Reported | 2,251 | 1,533 | 3,193 | 6,977 |
| | | Expected | 3,373 (3,197 – 3,538) | 5,203 (4,793 – 5,598) | 7,512 (6,809 – 8,263) | 16,088 (14,799 – 17,400) |
| | | Averted | 1,122 (946 – 1,287) | 3,670 (3,260 – 4,065) | 4,319 (3,616 – 5,070) | 9,111 (7,822 – 10,423) |
| | Per variant Month | Reported | 450 | 256 | 246 | 291 |
| | | Expected | 675 (639 - 708) | 867 (799 - 933) | 578 (524 - 636) | 670 (617 - 725) |
| | | Averted | 224 (189 - 257) | 612 (543 - 678) | 332 (278 - 390) | 380 (326 - 434) |
| Total | Cumulatively | Reported | 3,693 | 2,663 | 4,677 | 11,033 |
| | | Expected | 5,390 (5,109 – 5,640) | 8,918 (8,178 – 9,633) | 10,797 (9,658 – 12,024) | 25,105 (22,944 – 27,297) |
| | | Averted | 1,697 (1,416 – 1,947) | 6,255 (5,515 – 6,970) | 6,120 (4,981 – 7,347) | 14,072 (11,911 – 16,264) |
| | Per variant Month | Reported | 739 | 444 | 360 | 460 |
| | | Expected | 1,078 (1,022 – 1,128) | 1,486 (1,363 – 1,605) | 831 (743 - 925) | 1,046 (956 - 1137) |
| | | Averted | 339 (283 - 389) | 1,043 (919 – 1,162) | 471 (383 - 565) | 586 (496 - 678) |

**Supplement 5B:** Mortality rates of reported and expected deaths*, and the percentage change between these, by age group and by VOC-period, between 26 January 2021 and 31 January 2023 in Belgium.

* using VE against mortality based on a modified proxy for COVID-19 death including reported symptoms or hospitalizations related to COVID-19

| Age groups in years | Mortality rates | Alpha | Delta | Omicron | Total |
|---|---|---|---|---|---|
| **65-79** | Reported | 4,1 | 2,6 | 1,6 | 2,4 |
| | Expected | 5,7 (5,4 - 5,9) | 8,6 (7,9 - 9,4) | 3,5 (3,0 - 4,0) | 5,3 (4,8 - 5,8) |
| | % change | 28,5 (24,6 - 31,4) | 69,6 (66,6 - 72,0) | 54,8 (47,8 - 60,5) | 55,1 (50,3 - 59,0) |
| **80+** | Reported | 15,2 | 8,9 | 8,4 | 9,9 |
| | Expected | 22,8 (21,6 - 23,9) | 30,3 (27,9 - 32,6) | 19,7 (17,9 - 21,6) | 23,0 (21,2 - 24,9) |
| | % change | 33,5 (29,8 - 36,6) | 70,6 (68,1 - 72,7) | 57,3 (52,9 - 61,1) | 56,8 (53,1 - 60,1) |
| **Total** | Reported | 7,3 | 4,4 | 3,5 | 4,6 |
| | Expected | 10,7 (10,2 - 11,2) | 14,8 (13,6 - 16,0) | 8,2 (7,3 - 9,1) | 10,4 (9,5 - 11,3) |
| | % change | 31,6 (27,8 - 34,6) | 70,1 (67,4 - 72,4) | 56,6 (51,5 - 61,0) | 56,1 (52,0 - 59,6) |
